# Optical Genome Mapping improves detection and streamlines analysis of structural variants in myeloid neoplasms

**DOI:** 10.1101/2024.01.02.24300691

**Authors:** Gordana Raca, Trilochan Sahoo, M. Anwar Iqbal, Teresa A. Smolarek, Brynn Levy, Barbara R. DuPont, Scott Ryall, Adrian M. Dubuc, Nikhil Shri Sahajpal, Jie Liu, Jun Liao, Zi-Xuan Wang, Aaron A. Stence, Natalya V. Guseva, James R. Broach, Cecelia Miller, Deqin Ma, James Blachly, Phillip Michaels, Ravindra Kolhe, Rashmi Kanagal-Shamanna

## Abstract

Accurate diagnosis and risk stratification of hematological malignancies require disease-specific laboratory testing procedures involving the use of hematopathology, flow cytometry, molecular, and cytogenetic testing. While individual laboratories develop unique workflows to accommodate volume, clinical needs, and staffing, cytogenetic laboratories generally require a multitude of targeted and genome-wide tests that detect clinically relevant aberrations in hematologic malignancies. Specifically, the frequent use of multiple FISH panels coupled with concurrent chromosome analysis, can be both labor, and resource intensive. Optical Genome Mapping (OGM) is a comprehensive cytogenetic solution for detecting structural variants with high resolution and increased accuracy for hematological malignancy subtypes at the DNA level without need of any cell culture regimens. A new software tool for analysis of OGM data called VIA (Variant Intelligence Applications), provides an integrative analysis, interpretation, and reporting solution for OGM and other datatypes. In this study, we performed retrospective review of 56 datasets, representing 10 unique myeloid cases to assess multi-user (technologist and laboratory director) analyses and classification. Interpretation and reporting of OGM results were 100% concordant between reviewers for four cases with negative results by standard of care (SOC) testing. For the other six cases, five pathognomonic gene fusions identified by SOC assays were unanimously reported as Tier 1A classification was unanimous for five sentinel gene fusion rearrangements identified by SOC. OGM also found additional structural variants of clinical relevance in five of the six cases that were not found by SOC methods. Leveraging automatic pre-classification of variants and a custom decision tree, the VIA software enabled complete analysis with a mean technologist review time (variant analysis and initial tier determination) of 30.7 minutes. The analysis, interpretation, and reporting workflow described in this pilot study provides a framework for standardized and streamlined reporting of clinically significant variant in myeloid malignancies using VIA.

## Introduction

Diagnosis and clinical management of hematological malignancies increasingly necessitates comprehensive cytogenenomic evaluation (Khoury et al. 2022, NHS Foundation Trust 2020, National Comprehensive Cancer Network 2009). However, current standard of care approaches (including karyotyping, fluorescent in-situ hybridization (FISH), and chromosomal microarray) offer relatively low resolution, rely on visual, and therefore subjective, interpretations, and require manual efforts from trained, highly specialized personnel. Moreover, no individual cytogenetic tests is presently sufficient to comprehensively assess both copy number and balanced structural aberrations at the necessary resolution. Multiple cytogenetic testing modalities are thus used sequentially or in tandem to rule in/out clinically relevant findings recommended by national and international guidelines (Khoury et al. 2022, NHS 2020, National Comprehensive Cancer Network 2009). While next generation sequencing (NGS) panel-based testing have rapidly emerged as an important component of laboratory testing for hematologic malignancies, its use is largely restricted to assessment of single nucleotide variants and small insertions and deletions. It should be noted that some panels do detect fusions and copy changes. Initially, Sequencing was heralded as a single test for genomic analyses of hematologic malignancies, however, this promise has not yet materialized. NGS, largely through whole genome sequencing (WGS), has been implemented (Haferlach et al. 2018, Duncavage et al., 2021) but, because of the complexity of bioinformatic analysis and high cost, the WGS-based approaches have not been widely adopted.

OGM is emerging as an alternative, and perhaps first tier, cytogenomic technique for hematologic malignancies, consolidating multiple legacy cytogenomic techniques. By combining optical analysis of ultra-high molecular weight DNA with the scalability of high throughput molecular approaches, OGM is an attractive assay to comprehensively analyze complex cancers. OGM uses fluorescent labels to map single molecules to a reference genome, detecting balanced structural rearrangements and copy number variants ranging from 5kbp to aneusomies and ploidy-level complexity. As with any new technology, the analysis workflow must be tailored to the needs of the clinical laboratory. Ease of validation and implementation as well as genome-wide resolution and sensitivity in variant calling (≥5 kbp at low allele fraction from a heterogeneous DNA sample) are central considerations related to use of OGM in routine testing. To date, OGM has been successfully validated as a laboratory developed test in many laboratories worldwide, under a diverse set of regulatory bodies (Yang et al. 2022, Rack et al. 2022, Sahajpal et al. 2022, Nilius-Eliliwi et al. 2023). However, for achieving full benefits from this new technology, in addition to wet-lab processing and raw data analysis, variant review, interpretation, and reporting have to be streamlined and tailored to the needs of clinical laboratories.

To provide guidance for the clinical implementation of OGM, the International Consortium for Optical Genome Mapping in Hematologic Malignancies has recently authored a framework for the clinical implementation of OGM in hematologic malignancies (Levy et al. 2024). This guidance has been assembled by a consortium of early adopters of OGM, who have successfully implemented the technology in their clinical laboratories. Among other practical recommendations, the International Consortium manuscript addresses interpretation and classification of OGM variants; it expands upon the existing scheme for classification of cancer variants into tiers of clinical significance, updating them from application in chromosomal microarray (Mikhail et al. 2019) and proposing OGM specific definitions and changes.

In the current study, we extended the framework for laboratory implementation of OGM proposed by the International Consortium with a procedure for analysis and reporting of OGM data using the Variant Intelligence Applications (VIA^TM^) software to hematologic malignancy datasets. VIA (previously branded NxClinical) enables fast, automated, systematic, decision-tree based variant analysis and reporting of OGM data leveraging the well-established tools employed for years for clinical analysis of chromosomal microarray and next generation sequencing data (Chaubey et al. 2020). Here, we report a streamlined, laboratory-ready analysis workflow for OGM data in VIA, focused on myeloid neoplasms. The workflow was tested on 10 retrospective samples which were analyzed 5-6 times each by independent technologist-director teams to assess efficiency and reproducibility. Technologists timed the review process starting with the output of the SV calling algorithm through variant selection and pre-classification. After technologist review, variants were reviewed by a clinical laboratory director and given final classifications and included into VIA generated reports.

## Methods

### Cohort Information

Ten unique cases of myeloid malignancies [comprising nine cases of acute myeloid leukemia (AML) and one myelodysplastic syndrome/neoplasm (MDS)] were selected for this analysis workflow multi-technologist, multi-director trail. These cases were previously assessed by karyotype and FISH, in most cases, and also processed and classified using only Bionano Access and Bionano Solve software versions 1.7/3.7 (Pang et al. 2023).

### Data Collection

A single laboratory site received samples as aliquots of bone marrow aspirate or peripheral blood. Tissues were processed directly for genomic DNA (gDNA) isolation and labeling using Bionano reagent kits according to manufacturer’s instructions. Briefly, a 1.5 million cell volume equivalent was carried forward into lysis and gDNA binding reactions in the presence of a paramagnetic disk. Bound gDNA was washed and eluted in buffer, with 750ng carried forward into direct label and stain (DLS) reactions. Labeled DNA was then pipetted into a Saphyr G2.3. chip flowcell, Chips were placed in Saphyr Systems (part number 90067) and were run with an experimental design targeting 1500 Gbp data per flowcell. Datasets underwent quality control using the Molecule Quality Report (targeting (N50≥150) of ≥230 kbp, map rate ≥ 70% to GRCh38, and estimated effective coverage ≥340×). Files were then transferred into a central instance of Bionano Access for collaborative data management and analysis.

### Analysis and Processing through Access and VIA

The Rare Variant Analysis in Bionano Solve v3.7 and 3.8 was run using GRCh38 (hereafter: hg38) as reference and remaining default settings. OGM datasets used in this study were previously technically compared to reported standard of care (SOC) results by an unblinded individual not involved in case review, as analyzed in Bionano Solve version 3.7 and described in Pang et al. 2023. Each Rare Variant Analysis Informatics Report here was checked to confirm acceptable data quality in terms of effective coverage of the reference (target ≥300× for ≥5% VAF sensitivity) and CNV statistics (targets ≤20% above expected coefficients of variation, ≤0.25 correlation with label density). Certain datasets falling short of these QC targets have previously shown full technical concordance to SOC, and hence were accepted for this pilot study.

The previous Bionano Access interpretive workflow and analyst curation time data are presented here for comparative purposes. Briefly, it relies entirely upon Bionano Access and the application of custom filtering steps and region-specific features (BED files) for sequential curation of variants. Curated variants are then stepwise evaluated based on Access-provided annotations, then classified with built-in ACMG classifications. This analysis scheme relies on at least two reviewers’ analysis—one or more by an analyst (laboratory technologist), and once by a supervisor (credentialed laboratory director).

The data flow and replicate analysis scheme for this study are summarized in Figure 1. Datasets reanalyzed in Solve 3.8 were first reassigned to a newly available hg38_DLE1_0kb_0labels_masked_YPARs.cmap reference, and molecule quality reports were generated. The alignment then analyzed with the rare variant analysis with default settings including SV annotation against an OGM controls database of 394 phenotypically healthy individuals. Zipped pipeline results were then transferred into VIA with these SV results encoded in ogm.vcf files. Accompanying ogm.bam, and ogm.bai outputs were assigned the appropriate hg38 300× masked multiscale reference for further processing through the SNP FASST3 algorithm. This analytical step computes copy number and B-allele frequency profiles and provides subsequent CNV and allelic variant (allelic imbalance or absence of heterozygosity, hereafter AI/AOH) segments. Datasets were configured to preload AML or MDS guideline annotations as appropriate.

**Figure 1.**
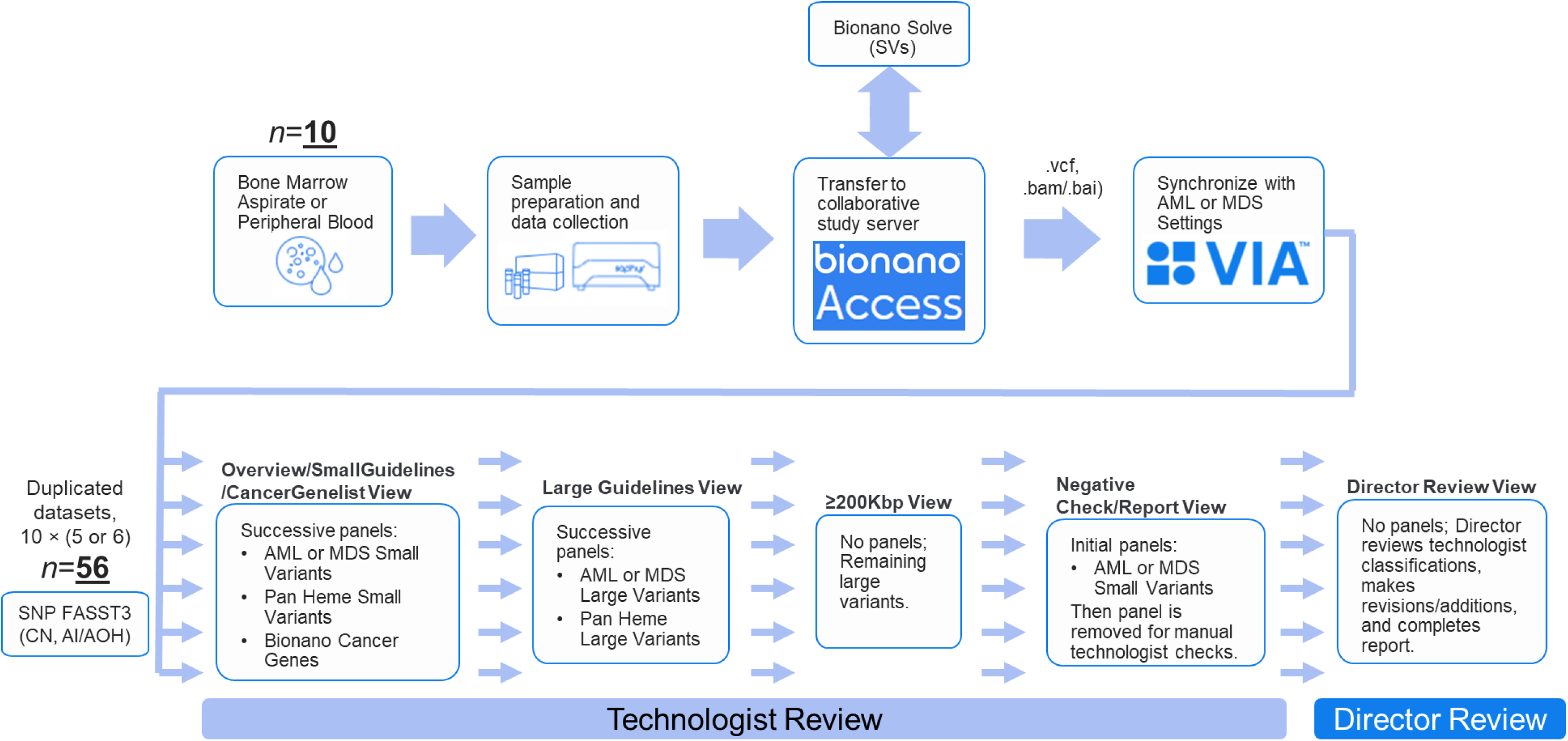
Data Flow and Replicate Analysis Framework. Ten bone marrow aspirate or blood samples from individuals with myeloid indications were prepared for OGM data collection on the Saphyr System. Datasets targeting ≥300× effective coverage of GRCh38 were transferred to a shared study server and analyzed using Solve 3.8 Rare Variant Analysis for .vcf (SVs), .bam/.bai outputs. These files were synced to a shared VIA server and assigned an appropriate 300× multiscale reference, duplicated into multiple copies for replicate review, and processed for CN and BAF data using the SNP FASST3 algorithm. Each dataset then underwent five or six reviews, proceeding in two stages: Technologist review, which consists of panel and filter application in the Overview/SmallGuidelines/CancerGenelist, Large Guidelines, ≥200Kbp, and Negative Check/Report Views; and a subsequent Director review for final classification and summary report generation.

### VIA Configuration for the study

The ten datasets used in this study were copied into at least six replicate VIA cases (for a multi-reviewer ring trial design) within partitioned VIA Sample Types. A suite of custom configurations in the VIA Sample Types was applied to tailor VIA’s interface to specific needs for this pilot study. These selections are reported in detail in the supplemental material. Briefly, five custom View settings (Table S1) were devised, which are abstracted filters, layouts, and panel selections that can be copied and furnished to multiple reviewers without each having to apply the changes individually.

Custom variant classifications were also devised (Table S2) to better supply reviewers with detailed options to record their interpretations. These custom classifications include:

- *Solve CNV Mask Overlap*, which was substituted for *Artifact* in the decision tree for copy number variants overlapping masked hg38 regions as described here: https://bionano.com/wp-content/uploads/2023/08/CG-30110-Bionano-Solve-Theory-of-Operation-Structural-Variant-Calling.pdf

Three custom classifications for minor variants concomitant with a major tiered variant:

- *AI of a CNV* – allelic imbalance (AI/AOH) resulting from a copy number variant. For example, an absence of heterozygosity resulting from a monosomy.
- *CNV of an SV* – copy number variant associated with a structural variant. For example, a gain resulting from a tandem duplication.
- *SV of an SV* – a structural variant resulting from a more primary SV. For example, a reciprocal breakpoint partner in a balanced translocation, whereby the primary breakpoint supports a gene fusion fulfilling a Tier 1A guideline variant (Figure 3).

And,

- *Redundant*, which was used for any variant which is an uninformative copy of a different relevant variant.

### Automated Variant Analysis, Technologist, and Director Review

Each replicate copy of the ten datasets underwent independent, blinded technologist and director reviews. Datasets were loaded in VIA by the technologist for preliminary review using a standard procedure devised for this study. Using the Overview/Small Guidelines/CancerGeneList view, the technologist first confirmed that the sex chromosome copy state in the whole genome copy number track matched the sex given in the clinical indication, to identify obvious sample mix up. With the copy number and B-allele frequency tracks loaded, the technologist then checked whether the diploid baseline calculated by the copy number algorithm was plausible given the underlying B-allele frequency datapoints. If a dataset was of a complex ploidy and the algorithm miscalculated diploid regions, the ploidy could be manually reset to a custom diploid region of the technologist’s choosing. Copy number data and AI/AOH segments would then be recalculated.

Next, the technologist checked CNV and AI/AOH segments for plausibility, and potential to consolidate adjacent fragments into merged segments at a final call level quality control step. VIA software enables users to interactively modify segment boundaries for CNV and AI/AOH calls with contradictory B-allele frequency data points or interstitial structural variant allele fraction evidence. Such segments were modified to new boundaries to exclude any contradictory datapoints, effectively shortening or eliminating segments likely to be spurious. Technologists were also empowered to manually assign CNV or AI/AOH segments not already called, if supported visually and/or contextually.

With the segmental calls checked for plausibility and accuracy, the technologist began a timed variant curation and interpretation process, using the custom views, panels and filters built into VIA (Figure 2). Each technologist review began with the Overview/Small Guidelines/CancerGeneList view loaded in VIA, and then sequentially annotation tracks (AML or MDS Small Variants, then Pan Heme Small Variants, then Bionano Cancer Genes) to serve as filters presenting variants to review. Any variants meeting basic quality control thresholds at 0% frequency in the OGM specific controls database that overlapped these panel targets, were prioritized for first review. Technologists loaded each variant under consideration into VIA’s tracks view, using genomic location and overlap with provided annotations (genes, guideline regions, database information i.e. DGV, etc.) to assess the importance and assign a technologist classification.

**Figure 2.**
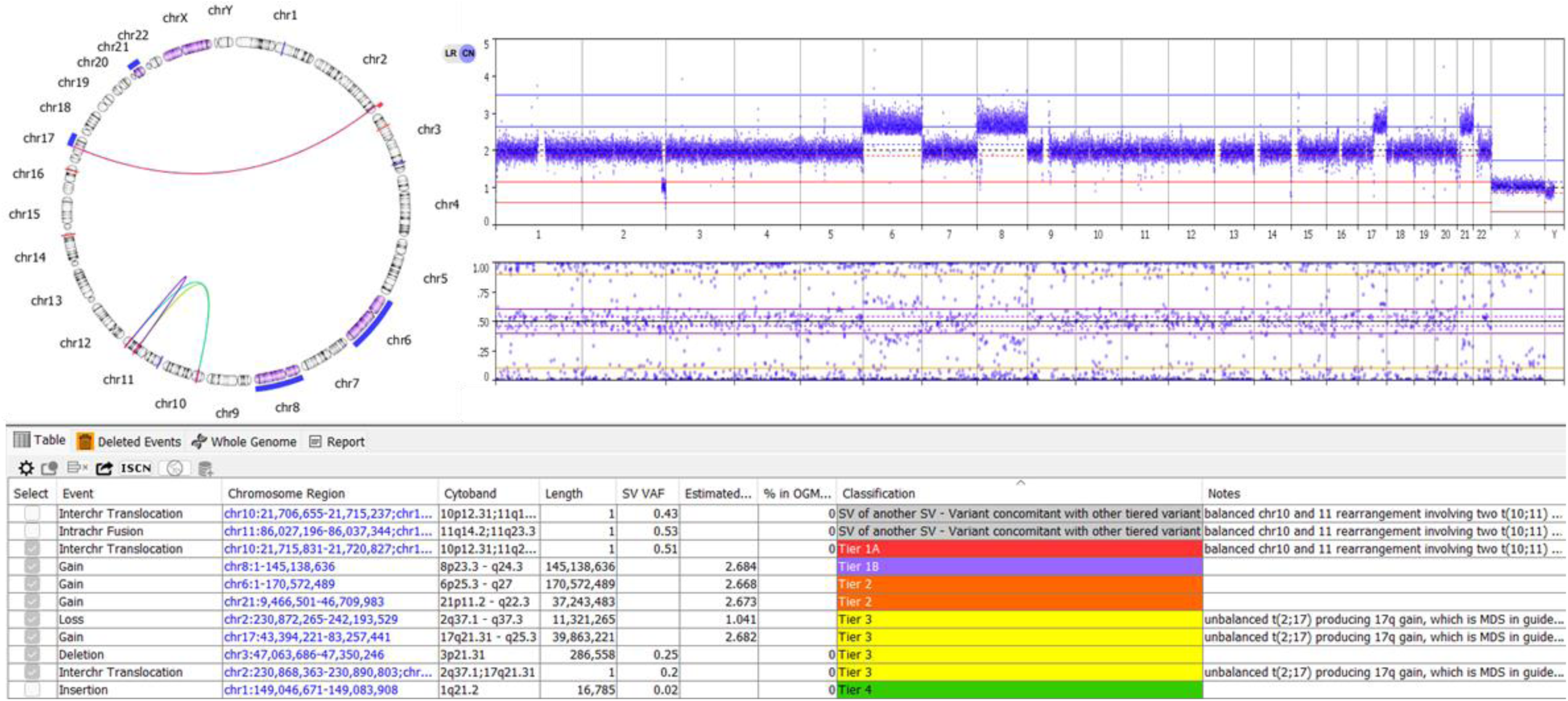
Case BNGOHM-0001008 loaded into VIA. The circos plot, whole genome (CN and BAF) plot, and variant table sorted by classification are shown.

**Figure 3.**
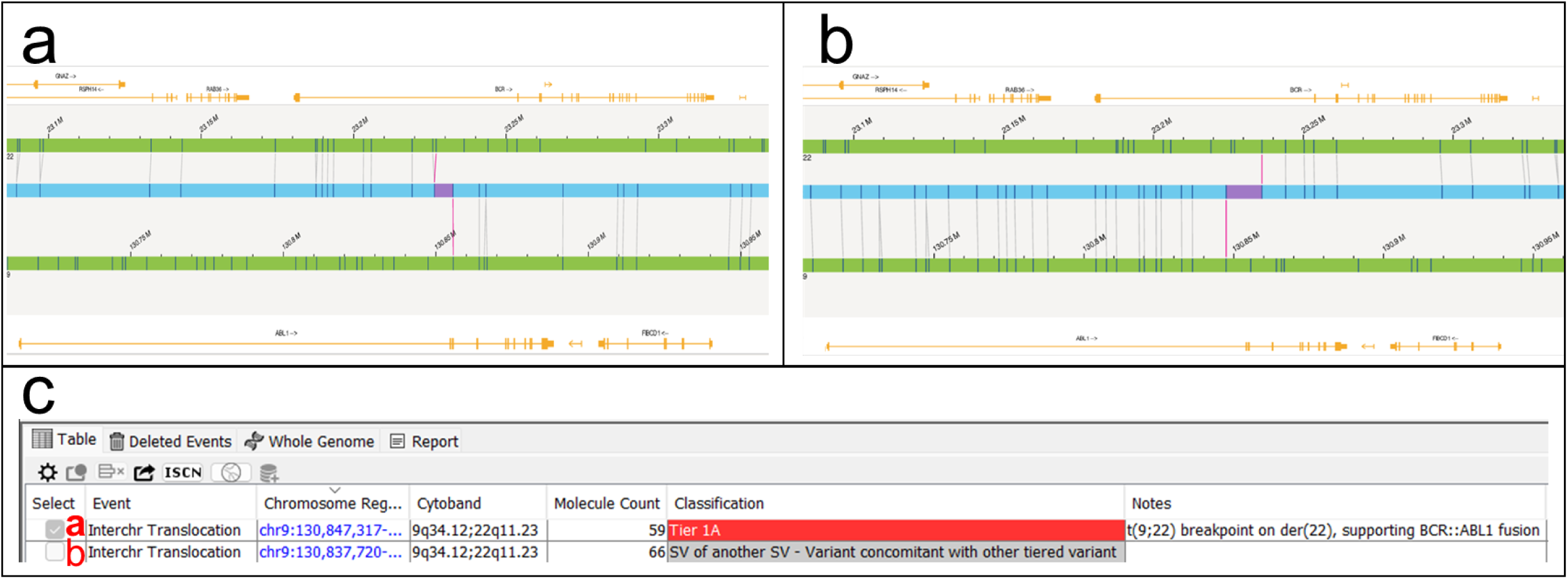
Case BNGOHM-0001054’s balanced t(9;22) and resulting SV calls, classifications. In top panels, Bionano Access shows center (blue) OGM consensus maps aligned partly to GRCh38 chromosome 22 (upper green track) and partly to chromosome 9 (lower green track). Reference gene annotations are shown in golden yellow. **a.** Map alignment showing the der(22) breakpoint, the configuration supporting a putative *BCR*::*ABL1* gene fusion. **b.** Separate map alignment shows the reciprocal der(9) breakpoint. **c.** Display and classification of both breakpoints in VIA: <u>Tier 1A</u> for the der(22) from panel a., and <u>SV of another SV – Variant concomitant with other tiered variant</u> for the der(9) from panel b.

This initial step identifies smaller variants with breakpoints directly overlapping a panel target of regions for consideration of Tier 1A classification. In a subsequent step, technologists applied a wider panel using target guideline regions across all hematological malignancies (“Pan Heme”). Variants overlapping these targets were considered for classifications. In a final step with this view loaded, the technologist applies a wide panel of cancer-associated genes generally (Bionano Cancer Genes) for consideration of overlapping variants.

With the full suite of small and gene-level panel targets interrogated, the technologist then loads the Large Guidelines view. This second view is designed to capture remaining large-scale abnormalities, such as aneuploidies and arm gains or losses. Two successive panels (AML or MDS Large Variants, then Pan Heme Large Variants) of disease-associated large abnormality regions are applied. This view captures virtually all remaining cytogenetic (>5Mb) abnormalities. The ≥200 kbp View is then loaded, which features further relaxed filters to present the technologist with all remaining variants ≥200 kbp. Any such large variant, absent in the OGM controls database, is considered for classification.

Finally in the Negative Check/Report View, all filters are released and the AML or MDS Small Variants panel is again loaded, to interrogate any remaining variant crossing a panel target. This step presents variants which may have been filtered out by quality score, yet nonetheless overlap a critical panel target. This panel is then removed to reveal all ≈1000-1200 variant calls, and relevant variants can be manually searched from this list if desired (not common). Finally, a negative check specific to three potentially difficult regions (the *IKZF1* gene, IGH region, and pseudoautosomal region 1) is performed in VIA. Once this is complete, the technologist notes the sex chromosomes of the dataset into the sample notes and records any desired information in the free text sample notes to guide the director’s review. The technologist then marked the review as “Tech 1 Complete” and stopped timing.

A director at laboratory director level at one of the affiliated institutions then began director review of the classifications. Directors first referred to the notes left by the technologist—the sex chromosomes of the dataset and sex provided in the clinical indication, as well as other notes about specific variant classification decisions. The director was then able to load any of the preceding views described, or the Director’s Review View, which focuses directly on those variants tiered by the technologist. The director sorted variants in the table by classification and/or audit log notes, then evaluated variants based on their expertise. The director was able to change the preliminary classification assigned by the technologist, and optionally leave notes providing rationale. Once the case variants were evaluated in director review, the director selected any variants deemed reportable (or, chose not to select variants for reporting). The director generated an ISCN summary in VIA for the report and entered an overall sample interpretation text summary. They then noted the cancer type (AML or MDS) and assigned a genome complexity (simple or complex) based on the number of reported variants. Finally, using the Target Guidelines feature in VIA, the director marked AML or MDS guideline variants as detected or not detected for the case. To complete review, director selected “Director review completed” and generated a case report using a Microsoft Word-compatible template and VIA’s built-in reporting features.

OGM variants selected as reportable, and their final classification tiers were considered for concordance analysis. Only the OGM findings reported in a plurality (two or more) of replicate reviews across technologist/director pairs were regarded for the study. Furthermore, we included only structural and copy number variants for analysis, and AI/AOH calls were excluded although VIA’s SNP FASST3 algorithm calculates B-allele frequency to make OGM AI/AOH calls.

## Results

A summary of all ten samples enrolled in the study with standard of care (SOC) results is provided in Table 1.

**Table 1.**
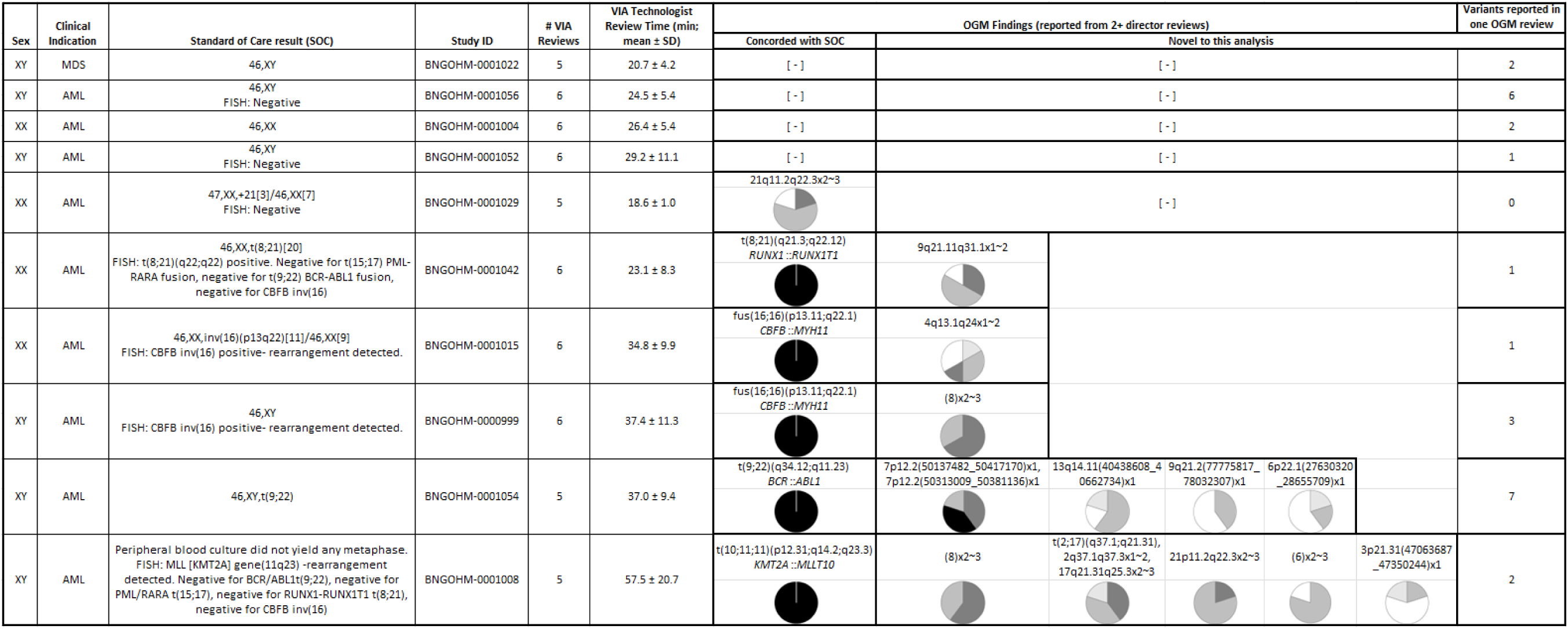
Summary of findings in the ten retrospective sample myeloid cohort. OGM findings are the same case variant(s) reported in two or more technologist/director paired reviews. Standard of care and OGM each report the same four cases as negative, while six contain one or more reported variants by SOC and OGM, tiered by n=5 or 6 director reviews as shown in the pie charts. The OGM workflow identified each SOC reported variant, and unanimously reported the sentinel *BCR*::*ABL1*, *CBFB*::*MYH11*, *KMT2A*::*MLLT10*, and *RUNX1*::*RUNX1T1* fusions as Tier 1A. Five cases contain variant(s) reported uniquely by the OGM workflow, which varied in frequency of reporting among directors and in tiering. Nine cases contained variants novel to OGM reported in only a single review, which are totaled in the rightmost column.

### Concordance analysis for SOC variant detection

After unblinding the SOC results, both SOC and OGM results identified at least one or more variants in six of 10 cases (56 replicates). The remaining four cases were negative by both SOC and OGM. All the results made and reported by SOC tests in this cohort were also identified by OGM via VIA (In one replicate, although +21 was identified by VIA, the variant was not selected for reporting).

### Concordance analysis for SOC variant reporting and classification

The single MDS case yielded a negative result for reportable variants in both SOC and OGM. Three of the nine AML cases were also negative, and six cases were positive for reportable finding(s). All classifications into tiers of clinical significance discussed in this study were hence decided in the context of samples with AML indications.

In five of six positive cases, the variant from SOC testing was classified as Tier 1A based on AML guidelines encoded into VIA. Each sentinel variant was unanimously reported and classified as Tier 1A in all OGM reviews (Figure 3). In the sixth case (BNGOHM-0001029), the SOC variant +21 was reported in four of five reviews with either Tier 1B or Tier 2 classification and omitted from reporting in one review. Reporting and classification decisions resulted in additional differences for non-sentinel AML variants, (none of which were not detected by SOC, they were uniquely identified by OGM, as discussed below).

### Novel findings detected by OGM only

There was a broad range of tier classifications in this category. OGM revealed additional variants which were not reported in the SOC analysis in five of six (87.5%) positive cases. In cases BNGOHM-0001015 and BNGOHM-0001042, OGM revealed large (>35Mbp) copy number losses in 4q and 9q, respectively, absent from the SOC reports. In case BNGOHM-0000999 OGM analysis showed trisomy 8, also absent from the reported SOC. The BNGOHM-0001015 4q13.1q24 loss was reported in four of six reviews, classifications ranging from Tier 1B to Tier 3; while the BNGOHM-0001042 9q21.11q31.1 loss was reported in five of six and classified as either Tier 1B or Tier 2.

Cases BNGOHM-0001008 and BNGOHM-0001054 showed more complex variants reported by multiple OGM reviewers. BNGOHM-0001054, an AML-M0 case, featured four interstitial deletions of 1.03 Mbp or less which would not be visible on a metaphase spread, and hence were not found in the SOC testing which for this case consisted solely of karyotype. Primary among these are two separate, overlapping deletions (68kbp and 280kbp) presumably comprising a bi-allelic loss of *IKZF1*. This event was reported in all five BNGOHM-0001054 reviews, with classifications ranging from Tier 1A to Tier 2. The remaining small deletions overlapped entries in the Bionano Cancer Gene List not specific to hematologic malignancy, and were variably reported and classified (Table 1).

BNGOHM-0001008 features a complex three breakpoint rearrangement of chromosomes 10 and 11, summarized here as t(10;11;11)(p12.31;q14.2;q23.3) was clearly shown by OGM to result in a *KMT2A*::*MLLT10* gene fusion, and was reported as a Tier 1A variant in all five reviews. The case featured an unbalanced t(2;17) which was reported in all OGM reviews and variably classified Tier 1B – Tier 3. Three chromosomal gains (+6, +8, and +21) were detected, reported in all but one instance of the +6, and classified as Tier 1B or Tier 2. A 3p21.31, 287 kbp interstitial deletion overlapping *SETD2* was reported in two of five reviews as Tier 2 or Tier 3. The SOC of BNGOHM-0001008 relies entirely on FISH results—which did identify an *MLL* rearrangement—as culture did not yield metaphase for karyotyping.

### Summary of Technologist Review timing in VIA

A summary of time to complete the technologist reviews can be found in Table 1. The speed of complete technologist case review averaged 25.2 minutes for the four negative cases, 28.5 minutes for four simple cases, and 47.3 minutes for the two OGM complex cases BNGOHM-0001008 and BNGOHM-0001054. Furthermore, different technologists had similar times for each case, as summarized in the Table 1 standard deviations. For example, case BNGOHM-0001042 was analyzed in 23.1 minutes on average with the fastest analysis in 14.0 minutes, and the slowest in 35.2 minutes. Other cases had more variation, for example case BNGOHM-0001008 (highest variation) had an average, fastest, and slowest times of 57.5, 31.4, and 86.9 minutes, respectively. Overall, the built-in workflow options with VIA and easy-to-master analysis workflow consistently kept the variant curation and initial technologist analysis time to a ≈20-45 minute average range for non-complex cases, depending on the number of findings. Reduced analysis time and labor was largely driven by the Views devised for this study, as opposed to managing multiple successive manual filter options. Also useful were point-and-click panel applications, and availability of layered browser tracks with which to analyze presented variants.

Nevertheless, VIA’s *View in Bionano Access* feature provided critical context to understand variants based on map alignments and label positions. Linking out to Access for alignment information was generally performed a handful of times for each case.

## Discussion

Cytogenetic testing is defined as the central genetic element of many national/international guidelines for the appropriate diagnosis, prognostication, or clinical assessment in many hematologic malignancies. Clinical standards rely on the use of karyotype and FISH to evaluate clinically relevant findings. In addition to these standards, microarrays and/or additional FISH studies are often necessary to clarify results. We created a data analysis paradigm to evaluate differences between current SOC approaches and OGM in respect to time, workload and reproducibility in interpretation and reporting of clinically significant variants.

OGM workflow automatically pre-classified tier 1A variants according to international guidelines including NCCN, WHO, NHS, resulting in the clear identification of *BCR*::*ABL1*, *CBFB*::*MYH11*, *RUNX1*::*RUNX1T1,* and *KMT2A::MLLT10* fusions. These were clearly and automatically flagged and presented to the technologist as putative tier 1A variants, which resulted in their unanimous identification and tier 1A classification.

Other variants were systematically and efficiently filtered by the review process devised for this study (Figure 1, Table S1), and were presented to the technologist for classification at tier 1B - tier 4, or other (non-tier) classifications (i.e. Table S2). This resulted in a range of 6 - 23 variants being manually classified in the cases presented here. Differences emerged between classifications assigned to non-tier 1A variants, in several cases resulting in differences among variants selected for reporting. Table 1 for example displays variable reporting of the same variants by blank (white) sections in the corresponding pie chart. In a few cases, the non-reported instance of the variant was not curated as a priority variant in the technologist review (the 9q21.11q31.1 loss in BNGOHM-0001042 for example). In others, the non-reported variant was evaluated and consciously omitted from reporting by the director (e.g. the (6)x2∼3 in BNGOHM-0001008).

The efficiency of this process is highlighted by the rapid processing of the datasets by the technologists. In this study, precise timing was tracked for every sample and replicate. Even with some variation in time for technologist review, it is highly similar and efficient. Review times in VIA were also compared to previous estimates by analysts at one affiliate site (Bionano Laboratories) for time needed when using Bionano Access 1.7 alone. VIA often yields a ≈50% time reduction while also adding several key features like customizable report generation and ability to store classified variants into a database and/or knowledgebase for assisting processing of future cases with same or similar variants.

Standard of care for the cohort was karyotype and FISH for six samples, karyotype alone for three samples, and FISH alone for one sample. OGM was able to detect all variants that SOC found, and reported, in a single assay, consistent with previously published reports (Yang et al. 2022, Levy et al. 2023, Pang et al. 2023). OGM further resolved more reportable findings in five of six non-negative cases (Table 1), including in two cases significantly more complex than karyotype was able to resolve (BNGOHM-0001054, karyotype 46,XY,t(9;22); and BNGOHM-0001008, no karyotype available as metaphase culture failed). Systematic improvement in resolving complex cancer genomes is a key benefit to applying OGM, while this workflow pilot shows that complex analyses can be completed efficiently alongside simple and negative cases.

Compared to current laboratory algorithms involving multiple FISH probes and karyotype, analysis using OGM with VIA offers improvements in analysis time and detection of all variant types at high resolution; it also adds the ability to report all results from a single assay. Labor shortages in cytogenetics has become a major issue in the clinical genetics and genomics laboratories and simplification of the testing algorithms through implementation of OGM offers an opportunity to alleviate this challenge.

## Conclusion

In conclusion, OGM analysis using Bionano Access and VIA offers a streamlined, highly efficient approach to interpret and report cytogenomic variants in hematological disorders. By its digital and highly objective nature, OGM and VIA have the potential to help standardize the analysis of variants and interpretation of their clinical significane, which to date is highly variable across the industry and internationally.

## Data Availability

Data will be made available upon reasonable request and in accordance with IRB protocols.

IRB and consent IRB-20212956 – Bionano Genomics Inc., San Diego, CA, USA

IRB-00007527 – University of Rochester Medical Center Office for Human Subject Protection

IRB-A-#00000150 (HAC IRB # 611298) - Medical College of Georgia, Augusta University, Augusta, GA, USA

## Acknowledgements

The authors wish to acknowledge Bionano clinical affairs team: Alka Chaubey, Alex Hastie, Andy Wing Chun Pang, Ileana Carrera, Alvin Yee, Jennifer Hauenstein, Ben Clifford, Stephanie Burke, Alex Chitsazan, Kelsea Chang, Simon Kim, James Yu, Mike Gallagher, Shuk Shukor, Alma Lastrella, Karl Hong, Beth Matthews, Sean Guy, Anusha Mylavarapu, Evelyn Crescini, Sheng-Wei Chang for their support with data management, training, conducting validation experiments and analysis.

---

**Table S1.**
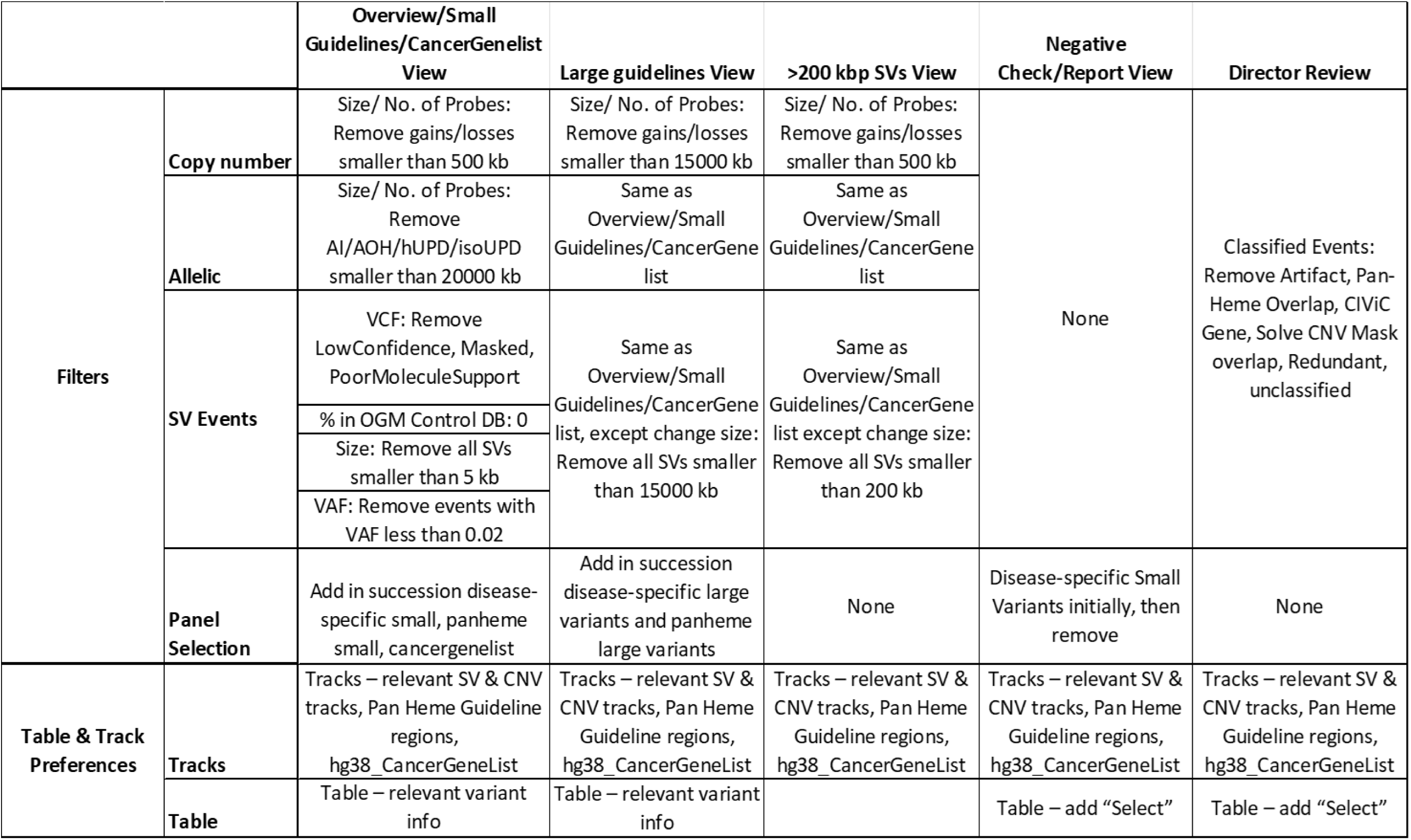
Filter, panel, and table & track preference settings applied in VIA for the five Views used in this study.

**Table S2.**
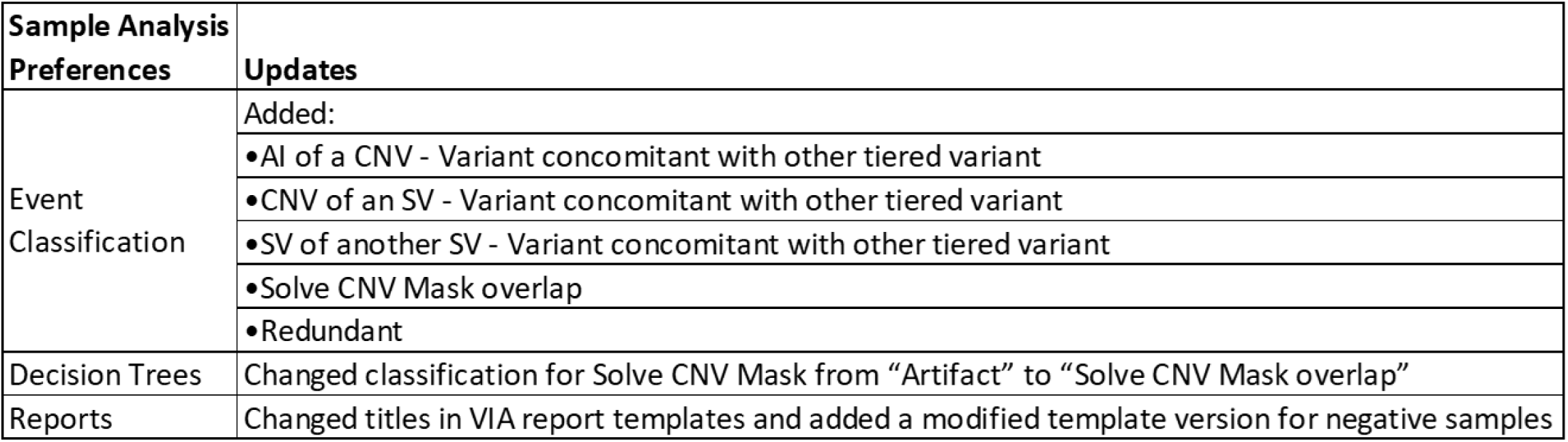
Custom VIA Sample Type configurations for this study.

| **Sample Analysis Preferences** | **Updates** |
| --- | --- |
| Event Classification | Added: |
|  | •AI of a CNV - Variant concomitant with other tiered variant |
|  | •CNV of an SV - Variant concomitant with other tiered variant |
|  | •SV of another SV - Variant concomitant with other tiered variant |
|  | •Solve CNV Mask overlap |
|  | •Redundant |
| Decision Trees | Changed classification for Solve CNV Mask from “Artifact” to “Solve CNV Mask overlap” |
| Reports | Changed titles in VIA report templates and added a modified template version for negative samples |

